# The MIND diet, brain transcriptomic alterations, and dementia

**DOI:** 10.1101/2023.06.12.23291263

**Authors:** Jun Li, Ana W. Capuano, Puja Agarwal, Zoe Arvanitakis, Yanling Wang, Philip L. De Jager, Julie A. Schneider, Shinya Tasaki, Katia de Paiva Lopes, Frank B. Hu, David A Bennett, Liming Liang, Francine Grodstein

**Author notes:** Address for Correspondence: Jun Li, MD, PhD, Address: 900 Commonwealth Ave. Boston, MA 02215.

## Abstract

Identifying novel mechanisms underlying dementia is critical to improving prevention and treatment. As an approach to mechanistic discovery, we investigated whether MIND diet (Mediterranean-DASH Diet Intervention for Neurodegenerative Delay), a consistent risk factor for dementia, is correlated with a specific profile of cortical gene expression, and whether such a transcriptomic profile is associated with dementia, in the Religious Orders Study (ROS) and Rush Memory and Aging Project (MAP). RNA sequencing (RNA-Seq) was conducted in postmortem dorsolateral prefrontal cortex tissue from 1,204 deceased participants; neuropsychological assessments were performed annually prior to death. In a subset of 482 participants, diet was assessed ~6 years before death using a validated food-frequency questionnaire; in these participants, using elastic net regression, we identified a transcriptomic profile, consisting of 50 genes, significantly correlated with MIND diet score (*P*=0.001). In multivariable analysis of the remaining 722 individuals, higher transcriptomic score of MIND diet was associated with slower annual rate of decline in global cognition (β=0.011 per standard deviation increment in transcriptomic profile score, *P*=0.003) and lower odds of dementia (odds ratio [OR] =0.76, *P*=0.0002). Cortical expression of several genes appeared to mediate the association between MIND diet and dementia, including *TCIM*, whose expression in inhibitory neurons and oligodendrocytes was associated with dementia in a subset of 424 individuals with single-nuclei RNA-seq data. In a secondary Mendelian randomization analysis, genetically predicted transcriptomic profile score was associated with dementia (OR=0.93, *P*=0.04). Our study suggests that associations between diet and cognitive health may involve brain molecular alterations at the transcriptomic level. Investigating brain molecular alterations related to diet may inform the identification of novel pathways underlying dementia.

## INTRODUCTION

Identifying pathways underlying dementia is critical to improved prevention and treatment, which are public health priorities. ^1^ Increasing epidemiological evidence supports the relationship between diet and dementia development. ^2^ Initial evidence from randomized clinical trials indicates that the Mediterranean diet and the Dietary Approaches to Stop Hypertension (DASH) may protect against cognitive decline. ^3–5^ The MIND diet (Mediterranean-DASH Diet Intervention for Neurodegenerative Delay) was specifically designed by tailoring the Mediterranean and DASH diets to emphasize foods and nutrients associated with dementia prevention. ^6, 7^ In multiple large-scale prospective cohorts, a higher MIND diet score is robustly associated with lower dementia risk and slower cognitive decline. ^6–12^

Understanding biological underpinnings of risk factors for dementia (such as diet) may inform the development of novel interventions for dementia prevention. Accumulating evidence from animal studies shows that calorie restriction or experimental feeding of specific nutrients or foods (such as polyphenols and polyunsaturated fatty acids that are abundant in the MIND diet) alters hippocampus and cortex expression of genes implicated in neurofunction and neuroinflammation, and improves brain plasticity and memory. ^13–15^ However, molecular mechanisms underlying the association between a healthy diet and dementia in humans are poorly studied.

We previously examined large scale RNA sequence (RNA-Seq) data from postmortem cortex tissue in two community-based, longitudinal cohorts – the Religious Orders Study (ROS) and Rush Memory and Aging Project (MAP); we found that the dorsolateral pre-frontal cortex (DLPFC) expression levels of several gene clusters – including genes broadly involved in metabolism, immune response, and synaptic transmission – were associated with dementia, cognitive decline, and Alzheimer’s disease (AD) pathology. ^16, 17^ Further, in ROSMAP, we previously found that cortical expression of specific genes might be on the pathway from dementia risk factors (such as neuroticism) to dementia. ^18^ Here, to extend this prior work, we leveraged the DLPFC RNA-Seq and cognitive data generated from 1,204 ROSMAP participants, and further integrated dietary data available in a subset of 482 MAP participants. We identified a cortical transcriptomic profile correlated with the MIND diet score, and tested whether such a transcriptomic profile was associated with cognitive trajectories and likelihood of dementia.

## SUBJECTS AND METHODS

Data and biospecimens for this study were drawn from ROS and MAP, two ongoing, prospective studies of aging and dementia, with a purposefully similar design. ROS started enrolling Catholic nuns, priests, and brothers from across the US in 1994. MAP began in 1997, enrolling residents from retirement communities and senior public housing from across the Chicago metropolitan area. At enrollment, participants were free of known dementia, ^19^ and agreed to annual clinical evaluations and organ donation at the time of death. ^20^ Structured neuropathologic examination was also performed after death. ^20^ At the time this study, over 3,700 participants were enrolled in ROSMAP and over 2,000 had died, with an overall follow-up rate in excess of 95% and an autopsy rate greater than 80%. Our study included the 1,204 ROSMAP participants with complete clinical and DLPFC RNA-Seq data^17, 21^ (**Figure 1**). The Institutional Review Board of Rush University Medical Center approved the study, and all participants signed an informed consent and anatomical gift act to donate tissue at the time of death.

**Figure 1.**
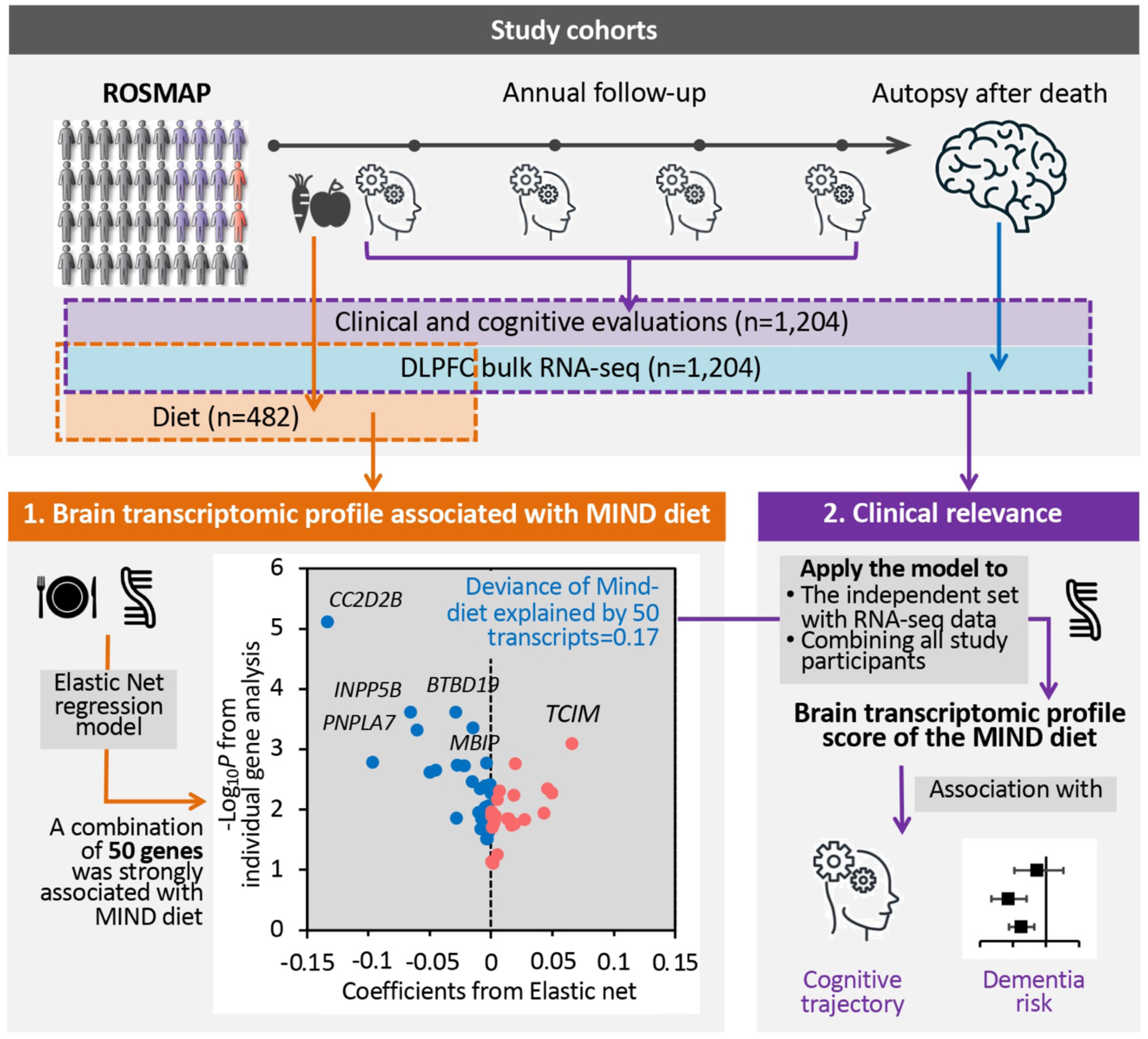
The flowchart of the study. We integrated dietary and clinical data, and bulk RNA sequencing (RNA-Seq) in dorsolateral prefrontal cortex (DLPFC) in ROSMAP. First, among 482 participants with dietary and RNA-Seq data, we applied an elastic net regression to regress the MIND diet score on all gene transcripts. The elastic net model identified a brain transcriptomic profile – constituting 50 genes – modestly but significantly associated with the MIND diet score in cross-validation analysis. Then, we applied this model to participants with RNA-Seq data (n=722 for the independent set, and n=1,204 combining all study participants) to calculate a brain transcriptomic profile score for the MIND diet and examined its association with cognitive trajectory (from baseline through death) and dementia status as of death.

### Assessment of Cognitive Health

Uniform structured clinical evaluations were performed at baseline and annually thereafter until death. ^20^ Briefly, detailed neuropsychological data from 17 harmonized tests across ROS and MAP were used to measure cognition in five cognitive domains. ^6, 20^ Raw scores from each test were first standardized based on means and standard deviations (SD) of the total group at baseline. A global composite score was calculated by averaging standardized scores across all tests. Individual trajectories of cognitive decline have been estimated among individuals with at least one follow-up cognitive assessment after baseline. ^22^ Briefly, mixed-effects models were applied to regress the global cognitive score on age, sex, and years of education as fixed effects (indicating average cognitive change over time in a group), together with a random intercept (indicating individual variation in cognition at baseline) and a random slope (indicating individual variation in cognitive changes over time). Person-specific slopes, adjusted for age, sex, education, were extracted from the models to quantitate individual cognitive decline rates. ^22^ This cognitive slope was used in our analysis to estimate cognitive trajectories.

At the time of death, all available clinical data were reviewed by a neurologist with expertise in dementia to determine the most likely cognitive diagnosis as of death, while blinded to postmortem data. ^19, 20, 23, 24^ Dementia was classified based on criteria of the joint working group of the National Institute of Neurological Communicative Disorders and Stroke and the Alzheimer’s Disease and Related Disorders Association, which require a history of cognitive decline with impairment in memory and at least one other cognitive domain. ^19, 23^ Mild cognitive impairment (MCI) was defined by the presence of cognitive impairment but no dementia and determined by clinicians as described in prior papers. ^19, 23^

### Dietary Assessments and the MIND Diet Score

Beginning in 2004, MAP participants were invited to complete a food frequency questionnaire (FFQ) at the time of their annual examinations, and 1,068 participants had FFQ data available at the time of analysis. ^6, 7^ The FFQ inquired about participants’ usual intake frequencies for over 144 food items in the past year and has been validated in older community residents. ^6, 7, 25^ The MIND diet score ranges from 0-15 (higher score represents healthier diet), summarizing 15 dietary components including 10 brain-healthy food groups (i.e., green leafy vegetables, other vegetables, nuts, berries, beans, whole grains, fish, poultry, olive oil, and wine) and 5 unhealthy food groups (i.e., red meats, butter and stick margarine, cheese, pastries/sweets, and fried/fast food). ^7^ In MAP, the score was highly stable over time in repeated FFQ assessments. ^6^ Thus, in this study, the MIND diet score was calculated based on the first FFQ to represent long-term dietary quality, as has been done in prior publications. ^6, 7^

### RNA Sequencing and data processing

For RNA-Seq of frozen DLPFC bulk tissues, laboratory procedures and data processing are described in detail in previous publications. ^16, 21, 26^ Briefly, the gray matter was separated from white matter and vasculature and was homogenized to extract RNA using standard commercial kits. Quality controlled RNA samples were sequenced using the Illumina HiSeq or NovaSeq platforms with an average sequencing depth of 50 million reads. ^21, 26^ In ROSMAP, bulk RNA-Seq were conducted by multiple projects, ^16, 21, 26^ and all available data were reprocessed and harmonized for combined use. ^26^ In data reprocessing, pair-end RNA-Seq data were aligned to GRCh38 reference genome (GENCODE release v27) using STAR v2.6, ^27^ and Picard was used to assess the quality of the aligned data. ^28^ Then, transcript raw counts were calculated by Kallisto (v0.46) ^29^ and aggregated at the gene level to obtain gene counts separately in mRNAs and pre-mRNAs. Samples were removed if total reads mapped were less than 5 million; had sex mismatch, SNP mismatch, or regional swap; were in a sequencing batch with only a few samples; or were identified as outliers based on multidimensional scaling analysis. ^26^ Genes with >10 reads in >50% of samples were retained. ^26^ Conditional quantile normalization was applied to account for bias from gene length and GC content; the adjusted gene counts matrix was then converted to log2-CPM (counts per million), followed by a quantile normalization using *voom* implemented in the R package *limma*. ^30^ Finally, a linear regression model was applied to remove major technical confounders (i.e., sequencing batch, post-mortem interval, RNA quality number, total spliced reads reported by STAR aligner, and metrics reported by Picard and Kallisto) while preserving regional differences. ^26^ In total, 17,255 quality-controlled and annotated genes were included in analyses.

In secondary analysis, we included single-nucleus RNA-Seq data from a subset of 424 ROSMAP individuals. Methods have been described elsewhere. ^31, 32^ Briefly, grey matter was processed in batches of 8 individuals; and 5000 nuclei from each batch were pooled, prepared, and sequenced using either Illumina HiSeqX at the Broad Institute’s Genomics Platform or Illumina NovaSeq 6000 at the New York Genome Center, with a target coverage of 1 million reads per channel. ^31, 32^ Data were first processed by the CellRanger software (v6.0.0; 10x Genomics) and quality controlled. Each nucleus was assigned back to its participants based on its genotypes. Cell type of nuclei were determined, and pseudo-bulk matrices were created by summing counts per individual. ^31, 32^ Within each cell type, genes with CPM > 1 in 80% of samples were retained and normalized using *tmm.voom*. Our analysis included expression of 32 genes from 7 cell types.

### Assessment of covariates and potential confounders

Information on age, sex, race, ethnicity, self-reported years of school education, and smoking history was collected from a structured interview administered at baseline. For analysis of MIND diet score, potential confounders including physical activity and body-mass index (BMI) were acquired from the annual visit when FFQs were administrated. ^6, 7^ Physical activity (hours per week) was measured based on self-reported minutes spent within the previous 2 weeks on five activities: walking for exercise, yard work, calisthenics, biking, and water exercise. BMI was calculated from weight and height measured by trained staff at clinical evaluations.

### Statistical Analysis

To identify a brain transcriptomic profile correlated with the MIND diet, we applied an elastic net regression – a variable selection model^33^ – to regress the MIND diet score on all 17,255 genes (all standardized to mean=0 and SD=1), in 482 individuals with both DLPFC RNA-Seq and FFQ data (**Figure 1**). The model lambda was chosen using a 10-fold cross-validation (CV) approach. From among the whole transcriptome, the elastic net model selected a group of genes whose weighted combination (i.e., a transcriptomic profile) showed the strongest correlation with MIND diet score while removing genes unlikely to be related. We evaluated model performance based on the fraction of (null) deviance explained and the mean cross-validated error. Further, to calculate the Pearson correlation between MIND diet score and its transcriptomic profile in the same 482 individuals without overfitting, we calculate an unbiased transcriptomic profile score for each individual using an external 10-fold CV approach, that is, to acquire the transcriptomic profile score for each 10% of the individuals, a prediction model was trained using the rest of the independent 90% of individuals. Associations between MIND diet score and individual genes were analyzed using linear regression, adjusting for age at death, sex, education, total energy intake, and years from the first FFQ to death. In sensitivity analyses, we further adjusted for BMI, physical activity, and smoking; and further excluded individuals with dementia at the time of FFQ. False discovery rate (FDR) was used to correct for multiple testing. Genes with FDR<0.05 were considered as statistically significant.

We then applied the elastic net model (trained in all 482 individuals) to the remaining independent set of 722 individuals with bulk DLPFC RNA-Seq data, to calculate the transcriptomic profile score for the MIND diet (**Figure 1**). In each individual, the transcriptomic profile score was calculated as the weighted sum of the selected genes with weights equal to the genes’ coefficients from the elastic net model. We analyzed associations between the transcriptomic profile score and (a) cognitive trajectories using linear regression and (b) final consensus diagnosis of dementia or MCI at death (*vs.* no cognitive impairment [NCI]) using multinomial logistic regression. Models were adjusted for age at death, sex, education, and study cohort. Considering the development of the transcriptomic profile in the first 482 individuals (i.e., those with diet data) is agnostic to participants’ cognitive health, to enhance statistical power, we secondarily applied the model to the 482 individuals with diet data and conducted joined analyses of all participants (n=1,204), to test associations between transcriptomic profile score and cognitive outcomes. We also analyzed associations between the DLPFC expression levels of the selected genes and cognitive outcomes in all 1204 participants. For genes showing FDR<0.05, we further assessed the associations between their cell type specific expression levels and the corresponding cognitive outcome in 424 individuals with single-nucleus RNA-Seq data.

As a simplified approach to control for confounding by cell composition in all study samples, we estimated proportions of neurons, astrocytes, microglia, oligodendrocytes, and endothelial cells, based on bulk RNA-seq data using a deconvolution digital sorting algorithm (DSA) implemented in the R package CellMix. ^34^ Two sets of cell type marker genes were used (200 top marker genes per cell type, ^35^ derived based on a human brain single-cell RNA-seq data, ^36^ and then separately, a cell-sorted RNA-seq data^37^). We conducted sensitivity analyses to examine whether cell compositions confounded associations between gene expressions and MIND diet score, and between the transcriptomic profile score and cognitive outcomes.

Among 444 individuals with FFQ, RNA-Seq data, and free of dementia at time of the FFQ, we verified previously reported associations between the MIND diet score (based on FFQ) and cognitive outcomes, ^6, 7^ after adjusting for age, sex, education, smoking, physical activity, BMI, and total energy intake. We then examined whether any selected genes in the transcriptomic profile mediated associations between MIND diet score and cognitive outcomes, using multiple mediation analysis implemented in R package *mma*. ^38^

To assist with interpretation of the potential temporal relationship between the transcriptomic profile score of MIND diet and dementia, we conducted secondary Mendelian randomization (MR) analysis using an inverse-variance weighted method, ^39^ based on genome-wide associations study (GWAS) summary data obtained separately for the transcriptomic profile score and AD. GWAS of the transcriptomic profile score was conducted in 670 independent set individuals with genetic data, using RVTESTS^40^, adjusting for sex, age at death, and the first 4 genetic principal components. Given the limited sample size, independent genetic variants (LD clumping with r^2^<0.1 in a 1000kbp window) with *P*<1×10^-6^ were used as the genetic instrumental variable (IV) for the transcriptomic profile score. To maximize statistical power, we acquired the summary statistics from a large-scale GWAS of clinically diagnosed AD (21,982 confirmed cases and 41,944 cognitively normal controls). ^41^ Independent genetic variants with *P*<5×10^-8^ were used as IV of AD.

All analyses, except for GWAS, were conducted using R version 4.2.1.

## RESULTS

A flow chart of our study and analytic populations is presented in **Figure 1**. Participants were largely female (68%), with average age at enrollment approximately 81 years old (**Table 1**). Mean age at death was 90 years; 525 had dementia and 285 had MCI as of death. The subgroup of 482 MAP participants with FFQ data was demographically comparable with the total study sample (**Table 1**); diet was assessed approximately 6 years before death and mean MIND diet score was 7.5 (SD=1.5).

**Table 1.** Characteristics of ROSMAP participants included in this study.

| **Variables** | **Subset of participants with dietary and RNA-Seq data**<br>**n=482** | **Independent set of participants with RNA-Seq data**<br>**n=722** | **All participants with RNA-Seq data**<br>**N=1204** |
| --- | --- | --- | --- |
| Female, n (%) | 340 (70.5%) | 479 (66.3%) | 819 (68.0%) |
| Education, years | 15.0 (12.0, 16.0) | 18.0 (15.0, 20.0) | 16.0 (13.0, 18.0) |
| Cognitive slope across study span * | 0.021 (-0.050, 0.060) | 0.005 (-0.065, 0.044) | 0.010 (-0.062, 0.050) |
| Age at study entry, years | 82.5 (6.0) | 79.7 (7.2) | 80.8 (6.9) |
| At the first FFQ: |  |  |  |
| Age, years | 84.7 (5.9) | - | - |
| MIND diet score | 7.5 (1.5) | - | - |
| Total energy intake | 1806.1 (554.0) | - | - |
| Ever smoking, n (%) | 193 (40.0%) | - | - |
| Body mass index, kg/m<sup>2</sup> | 26.5 (4.9) | - | - |
| Physical activity, hours/week | 2.2 (0.5, 4.0) | - | - |
| At the time of death: |  |  |  |
| Age, years | 90.8 (6.1) | 88.8 (6.7) | 89.6 (6.6) |
| MCI, n (%) | 124 (25.7%) | 161 (22.3%) | 285 (23.7%) |
| Dementia, n (%) | 183 (38.0%) | 342 (47.4%) | 525 (43.6%) |

Continuous variables were presented as mean (standard deviation) except for education years and cognitive slope, which were presented as median (25<sup>th</sup> and 75<sup>th</sup> percentiles) as they were not normally distributed. Categorical variables were presented as n (%).
\* Cognitive slope was estimated only in those with at least 2 cognitive assessments; n= 429 for participants with dietary and RNA-Seq data, n=668 for the independent set, and n=1,129 for all participants with RNA-Seq data.

### A DLPFC transcriptomic profile of the MIND diet

To identify a brain transcriptomic profile for the MIND diet, we applied an elastic net regression on all 17,255 genes simultaneously in 482 individuals with diet data. The model selected a combination of 50 genes showing the strongest association coefficients with MIND diet score, while robust to the effects of collinearity between genes (**Figure 2 and Supplementary Figure S1**). The model’s fraction of null deviance explained was 0.17, and the cross-validated r-square was 0.035. An unbiased transcriptomic profile score of the MIND diet calculated in the same 482 individuals using a 10-fold cross-validation approach (to avoid overfitting) was modestly but significantly correlated with MIND diet score (r= 0.15; *P*=0.001) (**Supplementary Figure S1**); this modest correlation was expected given the complex pathway from baseline diet to brain molecular changes identified postmortem. ^13, 14, 42^

**Figure 2.**
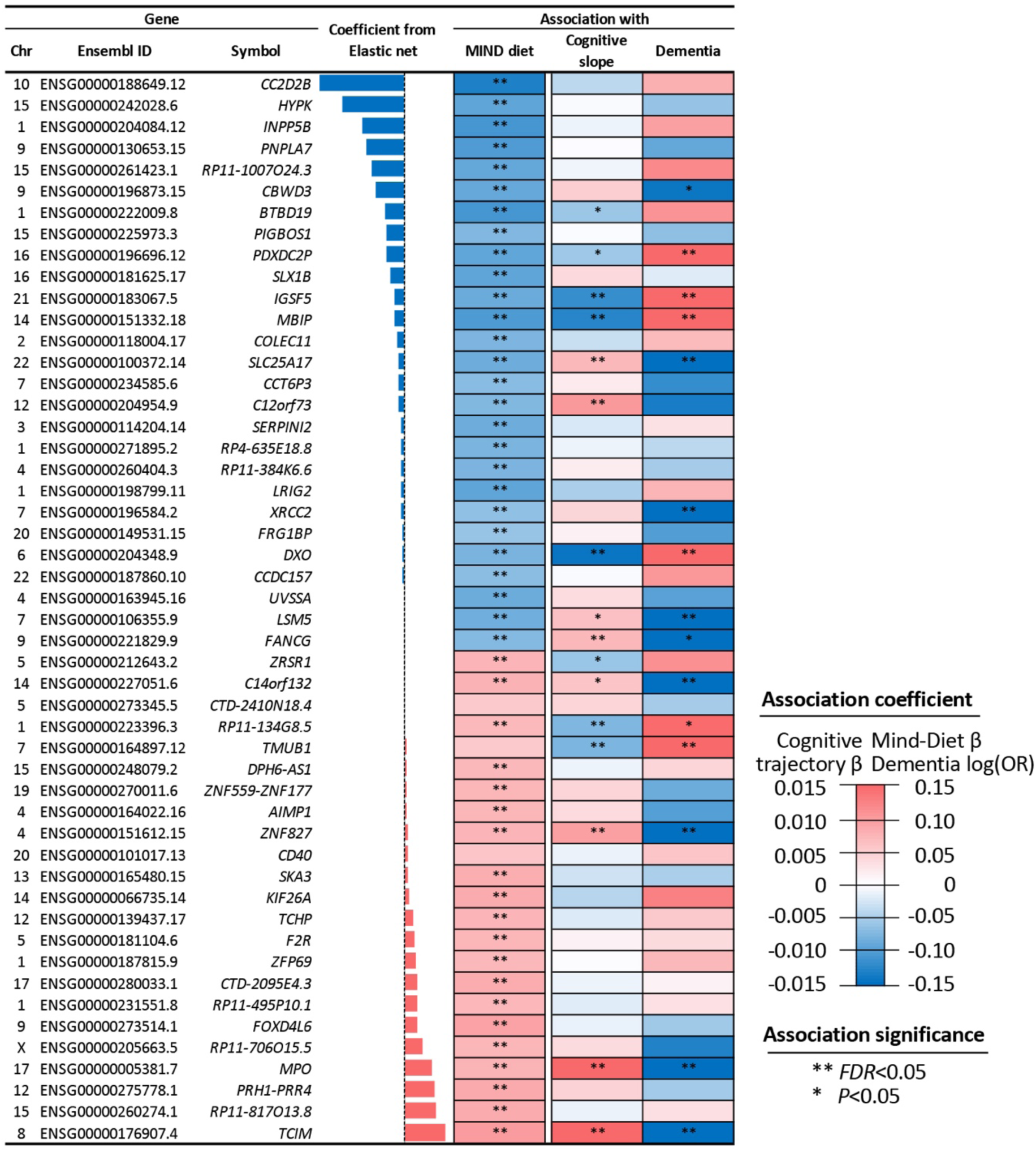
The 50 genes selected into the transcriptomic profile score of the MIND diet. The three columns indicate chromosomes, Ensembl ID, and symbol. For elastic net coefficients, blue and red bars indicate negative and positive coefficients, respectively, and bar heights indicate coefficient magnitudes. The three columns to the right are associations between each gene with MIND diet score (n=482), cognitive slope (n=1,129), and dementia (n=1,204). Colors indicate association directions, and color depths depict magnitudes (linear regression β for MIND diet score and cognitive slope; log [OR] for dementia). Blue color represents a negative β (i.e., a negative association) for MIND diet and cognitive decline, and an odds ratio below 1 (i.e., lower odds) for dementia. Red color represents a positive β for MIND diet and cognitive slope, and an above 1 odds ratio for dementia. Analyses were adjusted for age at death, sex, education, cohort, total energy intake, and years from first FFQ to death (only for MIND diet score).

Of these 50 selected genes, 23 were positively weighted in the transcriptomic profile (of which 21 genes were positively associated with MIND diet score at FDR<0.05 in individual gene analysis), and the other 27 genes were negatively weighted (all showed negative associations with MIND diet score at FDR<0.05) (**Figure 2**). Genes showing the strongest positive associations with MIND diet included *TCIM* and *MPO*, which have been related to educational attainment, and cognitive performance in depression in previous GWAS. ^43, 44^ Genes showing the strongest negative associations included *CC2D2B*, *PDXDC2P,* and *MBIP*, which are linked to AD, brain structure (e.g., white matter hyperintensities), and educational attainment; ^43, 45–47^ and *INPP5B* and *IGSF5*, which have been related to cardiovascular outcomes^48, 49^ in prior genetic studies (**Figure 2, Supplementary Table S1**). In sensitivity analyses, associations between MIND diet score and the 50 genes did not materially change after further adjusting for estimated cell type proportions, other lifestyle factors, or excluding individuals classified with dementia at baseline (**Supplementary Figure S2**).

### Gene expression and cognitive outcomes

We applied the 50-gene elastic net model to the remaining independent set of 722 individuals with RNA-Seq data, to acquire a transcriptomic profile score for the MIND diet and assessed its clinical relevance. In multivariable-adjusted analysis, a higher transcriptomic profile score was associated with slower rate of cognitive decline (β of cognitive slope per SD increment in transcriptomic profile score =0.011, *P*=3.5×10^-3^) and lower odds of dementia (OR=0.76 *vs.* NCI, *P*=0.003); but was not significantly associated with MCI (OR =0.89 *vs.* NCI, *P*=0.3) (**Figure 3**). A joint analysis including the 482 individuals with diet data and the independent set (total N=1,204) yielded highly consistent and more statistically significant findings (**Figure 3 and Supplementary Table S2)**. In sensitivity analysis, these associations did not materially change after further adjusting for the estimated cell type proportions (**Supplementary Table S3**).

**Figure 3.**
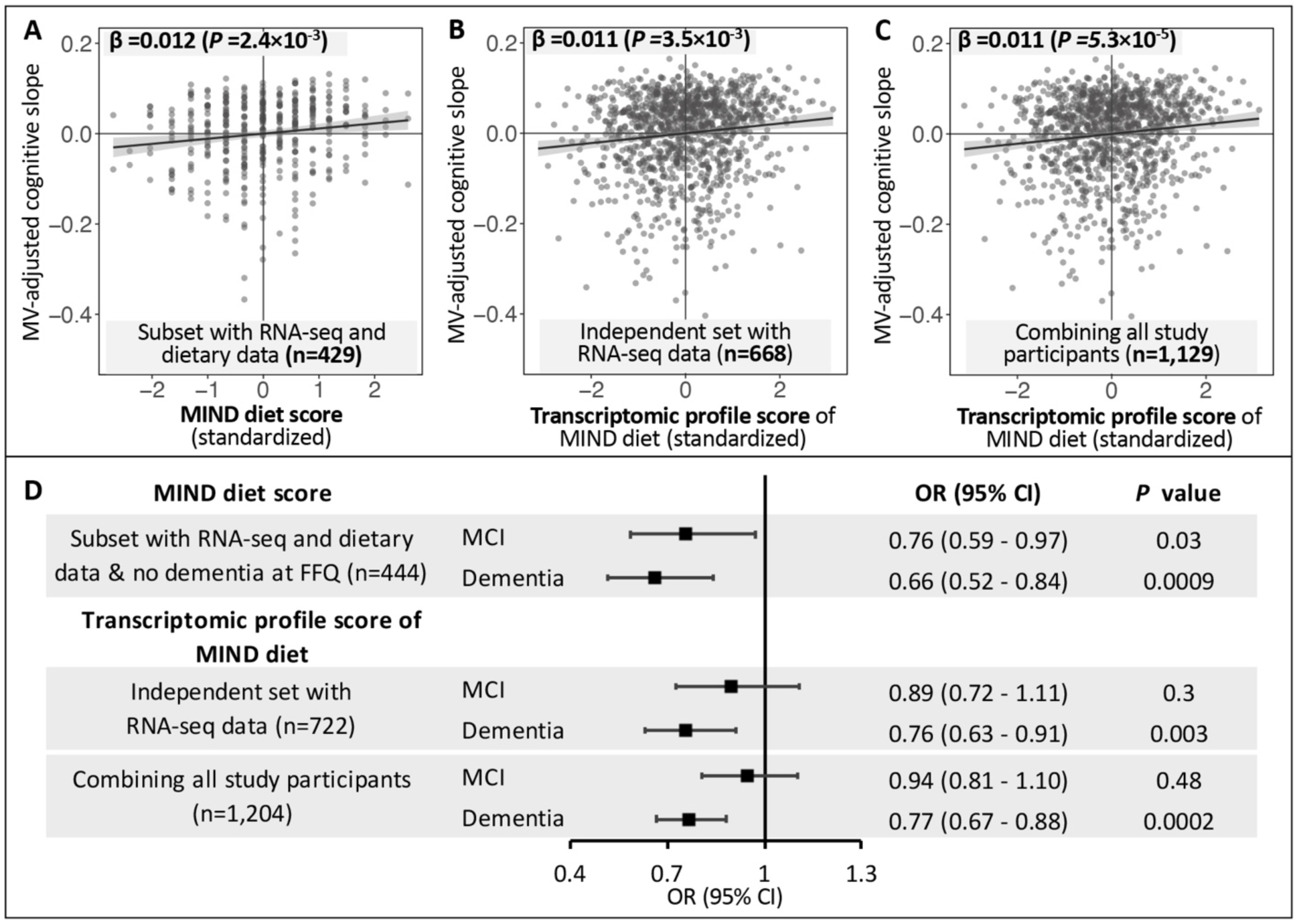
Associations of the MIND diet score and its cortical transcriptomic profile score with cognitive trajectory and dementia. The upper panel presents the associations with cognitive decline trajectory, for the MIND diet score (**A**), and for the transcriptomic profile score of the MIND diet in the independent set (**B**) and in the combined analysis of all participants (**C**). In the scatter plots, the solid lines are linear trend, and the grey areas are 95% confidence interval (CI). **D** presents the associations between the MIND diet score and its transcriptomic profile score with dementia and mild cognitive impairment (MCI) as of death (*vs.* no cognitive impairment). Black squares and grey lines indicate odds ratios (ORs) and 95% CI, respectively. Analyses of the MIND diet score were conducted in participants with dietary data and free of dementia at the time of the FFQ; and were adjusted for age, sex, years of education, BMI, physical activity, smoking, and total energy intake. Associations between the transcriptomic score were adjusted for age at death, sex, education years, and cohorts. Sample sizes were smaller in the upper panel *vs.* in D because some participants had only one cognitive assessment and were thus excluded from cognitive trajectory analysis.

Of the 50 genes constituting the transcriptomic profile, cortical expression levels of 11 genes were significantly associated with cognitive slope and 12 were associated with dementia (*FDR*<0.05), with highly consistent results from the independent set and the combined analysis of all participants (**Figure 2 and Supplementary Figure S3**). Notably, 4 individual genes (*TCIM*, *MPO*, *ZNF827*, and *C14orf132*) whose expression levels were associated with higher MIND diet scores, were also associated with lower odds of dementia and/or a slower rate of cognitive decline. Another 4 genes (namely *PDXDC2P*, *IGSF5*, *MBIP*, and *DXO*) whose expression levels were associated with lower MIND diet score were associated with worse cognitive outcomes (**Figure 2 and Supplementary Table S1**).

### MIND diet, gene expression, and cognitive outcomes

Among the 482 individuals with diet and RNA-Seq data, we confirmed that higher MIND diet score was significantly associated with slower rate of cognitive decline (β per SD increment in MIND score =0.012, *P*=0.002) and lower odds of dementia (OR=0.66, *P*=0.0009; *vs.* NCI) and MCI (OR =0.76, *P*=0.03; *vs.* NCI) as of death (**Figure 3**). Further, a multiple mediation analysis suggested that 3 genes potentially mediated the association between MIND diet and cognitive trajectory (*TCIM*, *DXO*, and *MPO*, accounting for 30% of the total association), and 5 genes mediated the association with dementia (*TCIM, DXO, MPO, PDXDC2P*, and *IGSF5*, accounting for 41% of the total association) (**Supplementary Figure S4**).

### Single nucleus RNA-Seq data

We then examined genes constituting the MIND diet transcriptomic profiles in single-nucleus RNA-Seq data (n=424; 32 genes out of 50 passed expression threshold in at least one cell type). We found that many of these genes were broadly expressed in all of the seven DLPFC cell types (e.g., *LSM5, C14orf132,* and *TMUB1*), with some restricted to specific cell types (e.g., *IGSF5* and *TCIM*) (**Supplementary Figure S5**). Further, of the 14 genes whose expression levels in bulk tissues were associated with cognitive trajectory and/or dementia, we observed consistent associations between 8 genes with cognitive outcomes in at least 1 of 7 brain cell types. For example, expressions of *TCIM* in inhibitory neurons and oligodendrocytes were associated with slower cognitive decline and lower odds of dementia, whereas expressions of *IGSF5* in astrocytes and excitatory neurons were associated with higher odds of dementia (**Supplementary Figure S6**).

### Secondary Mendelian randomization analysis

As RNA was sequenced in postmortem brain tissue, and thus it is possible that cognitive decline or dementia might cause alterations in gene expression rather than the reverse, we conducted a secondary MR analysis to assist interpretation of the potential temporal relationships between transcriptomic profile for the MIND diet and cognitive health. We found that the genetic IV of the transcriptomic profile score was marginally associated with lower odds of AD (OR =0.93 per SD increment in genetically inferred transcriptomic profile score, *P*=0.04), whereas genetically inferred AD was not associated with the transcriptomic profile score (*P*=0.11).

## DISCUSSION

Leveraging DLPFC RNA-Seq data in two longitudinal cohorts of aging, we identified a transcriptomic profile in the prefrontal cortex – consisting of 50 genes – correlated with the MIND diet score, and significantly associated with a slower rate of cognitive decline and lower odds of dementia. Our study investigating molecular pathways through which risk factors (e.g., diet) are associated with dementia showcases an approach to facilitate discovery of biological mechanisms in dementia, and to inform novel targets for dementia prevention.

Although evidence from clinical trials and prospective cohort studies supports that healthy diets can beneficially affect multiple cardiometabolic risk factors underlying dementia, such as lower systemic inflammation and risk of hypertension, diabetes, and stroke, ^50^ little research has explored molecular mechanisms in the brain linking diets to dementia in humans. Prior studies suggest a potential role of brain transcriptional regulation in dementia. ^16, 17, 26, 51, 52^ In ROSMAP, brain methylation levels of several CpGs were associated with burden of AD pathology. ^51^ Cortical expressions of an extensive list of genes were correlated with cognitive decline, dementia, and neuropathology; ^16, 17^ and a transcriptomic network module, consisting of 390 genes with diverse functions, was strongly associated with cognitive decline. ^16^ Recent studies further identified novel cortical proteomic modules associated with dementia, including some proteins potentially altered from the transcriptomic level. ^26, 52^ However, evidence on molecular changes linking diet to neurofunction has been mostly from animal studies. Feeding studies in animal models observed altered levels of hippocampal or cortical expressions of candidate genes related to metabolism, neuroinflammation, neurotransmission, and changes in brain plasticity and function. ^13–15^ In line with prior evidence, our study further suggests that in humans, behavioral risk factors such as diet are corrected with brain molecular profiles at the gene expression level.

In particular, our study highlighted several candidate genes of interest that are worth of future investigation. Notably, cortical expression of *TCIM* showed the strongest positive correlation with MIND diet and was a top mediator between MIND diet with slower cognitive decline and less dementia. *TCIM* encodes a *transcriptional and immune response regulator* which positively regulates the Wnt/β-catenin signaling pathway – a crucial pathway for neurogenesis, neuronal survival, and regulation of synaptic plasticity and blood-brain barrier integrity. ^53^ *TCIM* was previously identified as a genetic locus for educational attainment; ^43^ inhibitory neurons and oligodendrocyte – the cell types in which *TCIM* expression was associated with slower cognitive decline and lower odds of dementia in our study – have been functionally implicated in learning and memory. ^54, 55^ Our data suggested that healthy diets may upregulate cortical *TCIM* expression, and thus may help maintain cognition during aging.

In contrast, the MIND diet score was correlated with lower cortical expressions of *IGSF5 (Immunoglobulin Superfamily Member 5*), which appeared to be another mediator for the association between MIND diet and dementia. *IGSF5* encodes a junctional adhesion molecule, and its genetic variants have been associated with blood pressure, hypertension, and worse short-term memory. ^49, 56^ Interestingly, astrocytes – in which *IGSF5* expression showed the strongest association with higher odds of dementia in our study – appear to play a pivotal role in cerebral perfusion and brain blood flow. ^57^ In animal studies, “runner plasma”, collected from voluntarily running mice and infused into sedentary mice, downregulated brain *IGSF5* (among other genes) and correspondingly reduced neuroinflammation by targeting cerebrovasculature. ^58^ It is possible that healthy diets may exert similar effects as exercise on *IGSF5* expression, and function via vascular mechanisms.

More broadly, previous GWAS have linked other genes in our transcriptomic profile of MIND diet to inflammation (e.g., *DXO* and *MPO*) ^59^ and to cardiovascular disease (e.g., *INPP5B* and *ZNF827*). ^48, 60^ Inflammatory and vascular mechanisms have been associated with dementia risk, supporting the validity of our findings and further emphasizing the importance of targeting these pathways for dementia prevention.

Our study has important strengths. We uniquely leveraged a large sample of brain specimens with bulk RNA-Seq data (including a subset with single-nuclei RNA-Seq data) and clinical and cognitive assessments, in two longitudinal aging cohorts. The additional availability of dietary data in MAP further provided us an unprecedented opportunity to offer novel molecular insights into the association between MIND diet and dementia.

Our study has several limitations. First, our primarily analyses used RNA-seq in bulk DLPFC tissue that includes a mixture of cells. We estimated cell type proportions and showed that our primary results remained unchanged after adjusting for cell compositions; however, we cannot eliminate confounding by cell compositions. Although we examined single-nuclei RNA-Seq data in a sub-sample for selected genes, our statistical power was very limited, and we were unable to examine associations with diet due to little overlap with dietary data. Future studies investigating cell type-specific gene expressions in larger sample sizes will be important. Second, due to the use of postmortem samples, the temporal relationship between DLPFC gene expression and cognitive outcomes remains uncertain. Although our secondary MR analysis suggested that gene expression likely preceded dementia rather than the reverse, our statistical power for this analysis was limited due to the small sample size for GWAS of the brain transcriptomic profile score (although testing of the “reverse” directionality from dementia to the transcriptomic profile used 40 independent AD-associated variants as a strong instrument variable, and the results was null). Finally, although our analyses were adjusted for a wide range of potential confounders, unmeasured or residual confounding is possible. Future studies are warranted to validate our findings and elucidate the functionality and cell specificity of the identified genes in cognitive health.

In conclusion, we identified a brain transcriptomic profile correlated with the MIND diet, and this profile was significantly associated with slower cognitive decline and lower odds of dementia. Our study offered new insights into brain molecular mechanisms through which diet influences cognitive health and dementia, and these findings may inform the development of novel targets for dementia prevention.

## Data Availability

All phenotype data used in the study are available upon reasonable request to the Rush Alzheimer's Disease Center. All omics data used in the study are available online at AD Knowledge Portal (https://www.synapse.org/) upon reasonable request.

## Acknowledgements

We thank the study staff and participants of ROS and MAP for their essential contributions and donations of their brains to these projects.

## Authors’ contributions

Study design (J.L., A.W.C., Z.A., L.L, and F.G.); recruitment and data collection (P.A., Y.W., P.D.J., J.S., D.A.B.); bulk RNA-Seq and single-nuclei RNA-Seq data collection and/or processing (S.T., K.P.L., Y.W., P.D.J.); dietary data processing (P.A., and J.S.); data analysis (J.L., with supervision by A.W.C. and L.L and program review by A.W.C.); data interpretation (J.L., A.W.C., P.A., Z.A., P.D.J., K.P.L., F.B.H., D.A.B., L.L, and F.G.); drafting the article (J.L.); critical revision (J.L., A.W.C., P.A., Z.A., Y.W., P.D.J., J.S., S.T., K.P.L., F.B.H., D.A.B., L.L., and F.G); obtaining of funding (P.D.J., J.S., D.A.B.). All authors contributed to commenting and revising and have approved the publication of the paper.

