## Supplementary Information for "The MIND diet, brain transcriptomic alterations, and dementia"

Table of content


| | |
|---|---|
| Supplementary Table S1 | Associations between the 50 genes selected into the transcriptomic profile of the MIND diet with the MIND diet score, cognitive trajectory, and cognitive status as of death. |
| Supplementary Table S2 | Associations of the transcriptomic profile score of the MIND diet with cognitive trajectory, and dementia and MCI as of death (vs. no cognitive decline). |
| Supplementary Table S3 | Associations of the transcriptomic profile score of the MIND diet with cognitive outcomes after adjusting for predicted cell type compositions in combined analysis of participants. |
| Supplementary Figure S1 | The performance of elastic net regression to identify a DLPFC transcriptomic profile of the MIND diet. |
| Supplementary Figure S2 | Sensitivity analysis for associations between the DLPFC expressions of the 50 selected genes with the MIND diet score. |
| Supplementary Figure S3 | Associations between the DLPFC expression levels of the 50 selected genes with cognitive trajectory (A) and dementia risk (B), comparing results from the independent set (y-axis) with those from the combined analysis of all participants (x-axis). |
| Supplementary Figure S4 | Mediation effects of selected genes in the associations between the MIND diet score and cognitive outcomes. |
| Supplementary Figure S5 | Expression levels of genes in the transcriptomic profile of MIND diet by cell types, based on DLPFC single-nuclei RNA-Seq data in 424 ROSMAP participants. |
| Supplementary Figure S6 | Associations between cell type-specific expression levels of selected genes with cognitive trajectory (A) and dementia (B), in 424 ROSMAP participants with DLPFC single-nuclei RNA-Seq and clinical data. |

**Supplementary Table S1. Associations between the 50 genes selected into the transcriptomic profile of the MIND diet with the MIND diet score, cognitive trajectory, and cognitive status as of death.**

| Chr. | Gene ID | symbol | Elastic net coefficients | Association with MIND diet score (n=482) * | | | Association with cognitive slope (n=1,129) [#] | | | Association with dementia (n=1,204) † | | | Association with MCI (n=1,204) † | | |
|---|---|---|---|---|---|---|---|---|---|---|---|---|---|---|---|
| | | | | Est | P | FDR | β | P | FDR | OR | P | FDR | OR | P | FDR |
| 10 | ENSG00000188649.12 | *CC2D2B* | -0.1335 | -0.1316 | 7.6E-06 | 3.8E-04 | -0.0038 | 0.16 | 0.33 | 1.08 | 0.25 | 0.42 | 0.95 | 0.49 | 0.91 |
| 15 | ENSG00000242028.6 | *HYPK* | -0.0966 | -0.0931 | 1.6E-03 | 8.5E-03 | -0.0002 | 0.95 | 0.98 | 0.94 | 0.39 | 0.55 | 0.95 | 0.52 | 0.91 |
| 1 | ENSG00000204084.12 | *INPP5B* | -0.0659 | -0.1088 | 2.4E-04 | 4.0E-03 | -0.0009 | 0.75 | 0.86 | 1.10 | 0.17 | 0.33 | 1.15 | 7.0E-02 | 0.36 |
| 9 | ENSG00000130653.15 | *PNPLA7* | -0.0602 | -0.1030 | 4.8E-04 | 4.8E-03 | 0.0001 | 0.98 | 0.98 | 0.91 | 0.20 | 0.35 | 0.99 | 0.91 | 0.93 |
| 15 | ENSG00000261423.1 | *RP11-1007O24.3* | -0.0499 | -0.0900 | 2.4E-03 | 9.2E-03 | -0.0007 | 0.78 | 0.87 | 1.13 | 9.0E-02 | 0.22 | 1.03 | 0.71 | 0.91 |
| 9 | ENSG00000196873.15 | *CBWD3* | -0.0451 | -0.0897 | 2.2E-03 | 9.1E-03 | 0.0047 | 8.6E-02 | 0.24 | 0.87 | 4.2E-02 | 0.14 | 0.96 | 0.65 | 0.91 |
| 1 | ENSG00000222009.8 | *BTBD19* | -0.0290 | -0.1088 | 2.4E-04 | 4.0E-03 | -0.0057 | 3.3E-02 | 0.12 | 1.11 | 0.12 | 0.27 | 1.04 | 0.63 | 0.91 |
| 15 | ENSG00000225973.3 | *PIGBOS1* | -0.0285 | -0.0730 | 1.4E-02 | 1.9E-02 | -0.0001 | 0.98 | 0.98 | 0.94 | 0.35 | 0.51 | 1.05 | 0.54 | 0.91 |
| 16 | ENSG00000196696.12 | *PDXDC2P* | -0.0275 | -0.0918 | 1.8E-03 | 8.5E-03 | -0.0056 | 4.1E-02 | 0.13 | 1.24 | 1.7E-03 | 9.6E-03 | 1.18 | 3.8E-02 | 0.36 |
| 16 | ENSG00000181625.17 | *SLX1B* | -0.0220 | -0.0925 | 1.9E-03 | 8.5E-03 | 0.0036 | 0.18 | 0.36 | 0.98 | 0.80 | 0.82 | 1.03 | 0.71 | 0.91 |
| 21 | ENSG00000183067.5 | *IGSF5* | -0.0156 | -0.0864 | 3.4E-03 | 1.2E-02 | -0.0117 | 1.4E-05 | 1.4E-04 | 1.40 | 1.6E-06 | 2.7E-05 | 1.16 | 5.8E-02 | 0.36 |
| 14 | ENSG00000151332.18 | *MBIP* | -0.0148 | -0.1034 | 4.4E-04 | 4.8E-03 | -0.0127 | 3.2E-06 | 4.0E-05 | 1.30 | 1.8E-04 | 1.5E-03 | 1.08 | 0.32 | 0.80 |
| 2 | ENSG00000118004.17 | *COLEC11* | -0.0100 | -0.0755 | 1.1E-02 | 1.9E-02 | -0.0031 | 0.25 | 0.44 | 1.07 | 0.33 | 0.51 | 0.96 | 0.60 | 0.91 |
| 22 | ENSG00000100372.14 | *SLC25A17* | -0.0090 | -0.0840 | 4.5E-03 | 1.3E-02 | 0.0070 | 9.7E-03 | 4.9E-02 | 0.80 | 1.1E-03 | 8.0E-03 | 0.88 | 0.11 | 0.50 |
| 7 | ENSG00000234585.6 | *CCT6P3* | -0.0087 | -0.0684 | 2.1E-02 | 2.3E-02 | 0.0019 | 0.48 | 0.73 | 0.89 | 8.9E-02 | 0.22 | 0.94 | 0.42 | 0.91 |
| 12 | ENSG00000204954.9 | *C12orf73* | -0.0078 | -0.0727 | 1.4E-02 | 1.9E-02 | 0.0102 | 1.7E-04 | 1.4E-03 | 0.87 | 5.2E-02 | 0.16 | 1.05 | 0.55 | 0.91 |
| 3 | ENSG00000114204.14 | *SERPINI2* | -0.0052 | -0.0848 | 4.0E-03 | 1.2E-02 | -0.0021 | 0.43 | 0.70 | 1.03 | 0.68 | 0.71 | 0.92 | 0.26 | 0.69 |
| 1 | ENSG00000271895.2 | *RP4-635E18.8* | -0.0046 | -0.0771 | 9.3E-03 | 1.9E-02 | -0.0009 | 0.75 | 0.86 | 0.96 | 0.58 | 0.66 | 0.91 | 0.22 | 0.69 |
| 4 | ENSG00000260404.3 | *RP11-384K6.6* | -0.0043 | -0.0761 | 1.0E-02 | 1.9E-02 | 0.0017 | 0.53 | 0.78 | 0.95 | 0.46 | 0.56 | 1.12 | 0.15 | 0.61 |
| 1 | ENSG00000198799.11 | *LRIG2* | -0.0037 | -0.0930 | 1.7E-03 | 8.5E-03 | -0.0048 | 8.0E-02 | 0.24 | 1.08 | 0.29 | 0.46 | 0.97 | 0.70 | 0.91 |
| 7 | ENSG00000196584.2 | *XRCC2* | -0.0036 | -0.0641 | 3.0E-02 | 3.2E-02 | 0.0040 | 0.14 | 0.31 | 0.80 | 1.7E-03 | 9.6E-03 | 0.87 | 7.1E-02 | 0.36 |
| 20 | ENSG00000149531.15 | *FRG1BP* | -0.0030 | -0.0600 | 3.0E-02 | 3.2E-02 | 0.0012 | 0.68 | 0.86 | 0.90 | 0.18 | 0.33 | 0.90 | 0.24 | 0.69 |
| 6 | ENSG00000204348.9 | *DXO* | -0.0026 | -0.0779 | 8.6E-03 | 1.8E-02 | -0.0144 | 1.0E-07 | 1.7E-06 | 1.38 | 6.1E-06 | 6.5E-05 | 0.98 | 0.84 | 0.93 |
| 22 | ENSG00000187860.10 | *CCDC157* | -0.0019 | -0.0674 | 2.3E-02 | 2.6E-02 | -0.0001 | 0.96 | 0.98 | 1.11 | 0.13 | 0.29 | 1.11 | 0.17 | 0.61 |
| 4 | ENSG00000163945.16 | *UVSSA* | -0.0006 | -0.0855 | 3.8E-03 | 1.2E-02 | 0.0033 | 0.22 | 0.39 | 0.91 | 0.17 | 0.33 | 1.08 | 0.36 | 0.85 |

| | | | | | | | | | | | | | | | |
|---|---|---|---|---|---|---|---|---|---|---|---|---|---|---|---|
| 7 | ENSG00000106355.9 | *LSM5* | -0.0005 | -0.0828 | 5.2E-03 | 1.3E-02 | 0.0061 | 2.4E-02 | 9.9E-02 | 0.82 | 4.4E-03 | 2.0E-02 | 1.01 | 0.88 | 0.93 |
| 9 | ENSG00000221829.9 | *FANCG* | -0.0004 | -0.0714 | 1.5E-02 | 1.9E-02 | 0.0070 | 1.1E-02 | 5.0E-02 | 0.84 | 1.5E-02 | 5.9E-02 | 1.01 | 0.86 | 0.93 |
| 5 | ENSG00000212643.2 | *ZRSR1* | 0.0004 | 0.0738 | 1.3E-02 | 1.9E-02 | -0.0058 | 3.2E-02 | 0.12 | 1.12 | 0.11 | 0.27 | 0.90 | 0.21 | 0.69 |
| 14 | ENSG00000227051.6 | *C14orf132* | 0.0004 | 0.0757 | 1.1E-02 | 1.9E-02 | 0.0057 | 3.9E-02 | 0.13 | 0.81 | 2.6E-03 | 1.3E-02 | 0.84 | 2.8E-02 | 0.36 |
| 5 | ENSG00000273345.5 | *CTD-2410N18.4* | 0.0004 | 0.0532 | 7.4E-02 | 7.5E-02 | 0.0040 | 0.14 | 0.31 | 0.95 | 0.45 | 0.56 | 0.95 | 0.54 | 0.91 |
| 1 | ENSG00000223396.3 | *RP11-134G8.5* | 0.0007 | 0.0692 | 2.0E-02 | 2.3E-02 | -0.0073 | 7.2E-03 | 4.0E-02 | 1.16 | 3.4E-02 | 0.12 | 0.96 | 0.65 | 0.91 |
| 7 | ENSG00000164897.12 | *TMUB1* | 0.0016 | 0.0526 | 7.7E-02 | 7.7E-02 | -0.0076 | 5.0E-03 | 3.1E-02 | 1.20 | 7.9E-03 | 3.3E-02 | 1.06 | 0.44 | 0.91 |
| 15 | ENSG00000248079.2 | *DPH6-AS1* | 0.0021 | 0.0699 | 1.7E-02 | 2.1E-02 | -0.0009 | 0.74 | 0.86 | 1.04 | 0.57 | 0.66 | 0.94 | 0.46 | 0.91 |
| 19 | ENSG00000270011.6 | *ZNF559-ZNF177* | 0.0026 | 0.0723 | 1.5E-02 | 1.9E-02 | 0.0040 | 0.14 | 0.31 | 0.92 | 0.22 | 0.38 | 0.96 | 0.61 | 0.91 |
| 4 | ENSG00000164022.16 | *AIMP1* | 0.0029 | 0.0723 | 1.5E-02 | 1.9E-02 | 0.0035 | 0.19 | 0.37 | 0.91 | 0.16 | 0.32 | 1.02 | 0.83 | 0.93 |
| 4 | ENSG00000151612.15 | *ZNF827* | 0.0034 | 0.0735 | 1.3E-02 | 1.9E-02 | 0.0094 | 4.9E-04 | 3.5E-03 | 0.73 | 6.5E-06 | 6.5E-05 | 0.86 | 6.0E-02 | 0.36 |
| 20 | ENSG00000101017.13 | *CD40* | 0.0048 | 0.0563 | 5.6E-02 | 5.8E-02 | -0.0010 | 0.72 | 0.86 | 1.06 | 0.42 | 0.56 | 1.02 | 0.80 | 0.93 |
| 13 | ENSG00000165480.15 | *SKA3* | 0.0052 | 0.0803 | 6.7E-03 | 1.5E-02 | -0.0027 | 0.32 | 0.54 | 0.95 | 0.49 | 0.58 | 0.90 | 0.16 | 0.61 |
| 14 | ENSG00000066735.14 | *KIF26A* | 0.0068 | 0.0835 | 4.9E-03 | 1.3E-02 | -0.0041 | 0.13 | 0.31 | 1.13 | 6.9E-02 | 0.19 | 1.00 | 1.00 | 1.00 |
| 12 | ENSG00000139437.17 | *TCHP* | 0.0133 | 0.0727 | 1.4E-02 | 1.9E-02 | -0.0019 | 0.49 | 0.73 | 1.05 | 0.44 | 0.56 | 1.10 | 0.25 | 0.69 |
| 5 | ENSG00000181104.6 | *F2R* | 0.0147 | 0.0720 | 1.4E-02 | 1.9E-02 | 0.0011 | 0.68 | 0.86 | 1.04 | 0.62 | 0.69 | 0.98 | 0.75 | 0.93 |
| 1 | ENSG00000187815.9 | *ZFP69* | 0.0170 | 0.0703 | 1.8E-02 | 2.2E-02 | 0.0002 | 0.95 | 0.98 | 1.07 | 0.31 | 0.49 | 1.19 | 2.9E-02 | 0.36 |
| 17 | ENSG00000280033.1 | *CTD-2095E4.3* | 0.0182 | 0.0822 | 5.7E-03 | 1.3E-02 | -0.0009 | 0.73 | 0.86 | 1.01 | 0.87 | 0.87 | 0.96 | 0.57 | 0.91 |
| 1 | ENSG00000231551.8 | *RP11-495P10.1* | 0.0191 | 0.0707 | 1.7E-02 | 2.1E-02 | -0.0016 | 0.54 | 0.78 | 1.03 | 0.68 | 0.71 | 1.02 | 0.82 | 0.93 |
| 9 | ENSG00000273514.1 | *FOXD4L6* | 0.0196 | 0.0921 | 1.7E-03 | 8.5E-03 | -0.0011 | 0.69 | 0.86 | 0.95 | 0.44 | 0.56 | 0.85 | 4.9E-02 | 0.36 |
| X | ENSG00000205663.5 | *RP11-706O15.5* | 0.0273 | 0.0720 | 1.5E-02 | 1.9E-02 | 0.0034 | 0.20 | 0.38 | 0.88 | 6.4E-02 | 0.19 | 0.95 | 0.56 | 0.91 |
| 17 | ENSG00000005381.7 | *MPO* | 0.0433 | 0.0754 | 1.1E-02 | 1.9E-02 | 0.0155 | 7.8E-09 | 2.0E-07 | 0.67 | 1.9E-08 | 4.7E-07 | 0.99 | 0.91 | 0.93 |
| 12 | ENSG00000275778.1 | *PRH1-PRR4* | 0.0459 | 0.0843 | 4.5E-03 | 1.3E-02 | 0.0043 | 0.11 | 0.29 | 0.95 | 0.45 | 0.56 | 1.01 | 0.91 | 0.93 |
| 15 | ENSG00000260274.1 | *RP11-817O13.8* | 0.0494 | 0.0828 | 5.3E-03 | 1.3E-02 | -0.0010 | 0.72 | 0.86 | 1.03 | 0.67 | 0.71 | 1.18 | 3.9E-02 | 0.36 |
| 8 | ENSG00000176907.4 | *TCIM* | 0.0656 | 0.0991 | 8.0E-04 | 6.7E-03 | 0.0169 | 2.9E-10 | 1.4E-08 | 0.65 | 3.1E-09 | 1.6E-07 | 0.80 | 5.3E-03 | 0.26 |

Expression levels of genes were inverse normal transformed before association analysis. To increase comparison, the MIND diet score was standardized before analysis. Multiple testing correction was conducted using false discovery rate (FDR) on the 50 genes tested.

* Linear regression was adjusted for age at death, sex, education, cohort, total energy intake, and years from the first FFQ to death.

[#] Linear regression was adjusted for age at death, sex, education, and cohort.

† Multinomial logistic regression was adjusted for age at death, sex, education, and cohort, with no cognitive impairment as reference.

**Supplementary Table S2. Associations of the transcriptomic profile score of the MIND diet with cognitive trajectory, and dementia and MCI as of death (*vs.* no cognitive decline).**

| | Association with cognitive slope | | | Association with dementia | | | Association with MCI | | |
|---|---|---|---|---|---|---|---|---|---|
| | n | β (95% CI) | *P* | case n | OR (95% CI) | *P* | case n | OR (95% CI) | *P* |
| **In the independent set** | | | | | | | | | |
| In all (n=722) | 668 | 0.011 (0.003 - 0.018) | 0.004 | 342 | 0.76 (0.63 - 0.91) | 0.003 | 161 | 0.89 (0.72 - 1.10) | 0.30 |
| In women (n=479) | 448 | 0.009 (0.000 - 0.018) | 0.05 | 242 | 0.76 (0.61 - 0.96) | 0.02 | 104 | 0.92 (0.71 - 1.21) | 0.56 |
| In men (n=243) | 220 | 0.013 (0.002 - 0.024) | 0.03 | 100 | 0.78 (0.57 - 1.07) | 0.12 | 57 | 0.87 (0.61 - 1.23) | 0.42 |
| **In combined analysis of all participants with RNA-Seq data** | | | | | | | | | |
| In all (n=1,204) | 1,129 | 0.011 (0.006 - 0.016) | 0.0001 | 525 | 0.77 (0.67 - 0.88) | 0.0002 | 285 | 0.94 (0.81 - 1.10) | 0.48 |
| In women (n=819) | 774 | 0.010 (0.003 - 0.017) | 0.004 | 370 | 0.76 (0.64 - 0.90) | 0.001 | 187 | 1.00 (0.82 - 1.21) | 1.00 |
| In men (n=385) | 355 | 0.012 (0.004 - 0.021) | 0.005 | 155 | 0.81 (0.63 - 1.03) | 0.09 | 98 | 0.85 (0.65 - 1.12) | 0.25 |

We calculated the associations of the transcriptomic profile score with cognitive slope using linear regression; associations with MCI and dementia (*vs.* no cognitive decline) using multinomial logistic regression, adjusting for age at death, sex, education years, and cohort. To enable a comparison of the association coefficients with those from the MIND diet score, the transcriptomic profile score was standardized before analysis.

The smaller sample size for cognitive slope analysis was because some participants had only one cognitive assessment during follow-up and were thus excluded from the analysis of cognitive trajectory.

For all analysis, the test for association differences between sexes were ≥0.35.

**Supplementary Table S3. Associations of the transcriptomic profile score of the MIND diet with cognitive outcomes after adjusting for predicted cell type compositions in combined analysis of participants.**

| **Analysis models** | **Association with cognitive slope** | | **Association with dementia** | | **Association with MCI** | |
|---|---|---|---|---|---|---|
| | **β (95% CI)** | ***P*** | **OR (95% CI)** | ***P*** | **OR (95% CI)** | ***P*** |
| Further adjust for cell subtype proportions estimated based on | | | | | | |
| 1. Cell-type specific gene markers derived from brain cell-sorted RNA-seq data | 0.011 (0.005 - 0.016) | 0.0001 | 0.76 (0.66 - 0.88) | 0.0002 | 0.96 (0.81 - 1.12) | 0.59 |
| 2. Cell-type specific gene markers derived from brain single-cell RNA-seq data | 0.010 (0.005 - 0.016) | 0.0001 | 0.77 (0.66 - 0.88) | 0.0003 | 0.96 (0.82 - 1.13) | 0.64 |

Cell subtype proportions (for neurons, astrocytes, microglia, oligodendrocytes, and endothelial cells) were calculated using a digital sorting algorithm (DSA) based on cell-type specific gene markers derived from human brain cell-sorted RNA-seq data ("Darmanis") or human brain single-cell RNA-seq data ("Zhang").

Associations between the transcriptomic profile score and cognitive slope were calculated using linear regression; associations between the transcriptomic profile score with MCI and dementia (*vs.* no cognitive decline) were calculated using multinomial logistic regression, adjusting for age at death, sex, education years, cohort, and estimated proportions of neurons, astrocytes, microglia, and oligodendrocytes (the proportion of endothelial cells was not adjusted due to it being dependent on proportions of other cell types).

**Supplementary Figure S1. The performance of elastic net regression to identify a DLPFC transcriptomic profile of the MIND diet. A.** The volcano plot for the transcriptome-wide association analysis of the MIND diet score, adjusting for age at death, sex, education years, cohort, total calorie intake, and years from the first FFQ to death. Dots represent each of the genes and the 50 genes selected by the elastic net regression were highlighted in red. This comparison demonstrated the elastic net model selected top genes associated with the MIND diet score while considering the collinearity between gene transcripts. **B.** We developed the transcriptomic profile of the MIND diet in 482 participants with dietary and RNA-Seq data. Due to the lack of an independent validation set (in which both diet and DLPFC RNA-Seq data were available), to demonstrate the Pearson correlation between the MIND diet and its transcriptomic profile score in the same 482 participants while avoiding overfitting, we calculated an unbiased transcriptomic profile score using a 10-fold cross-validation approach.

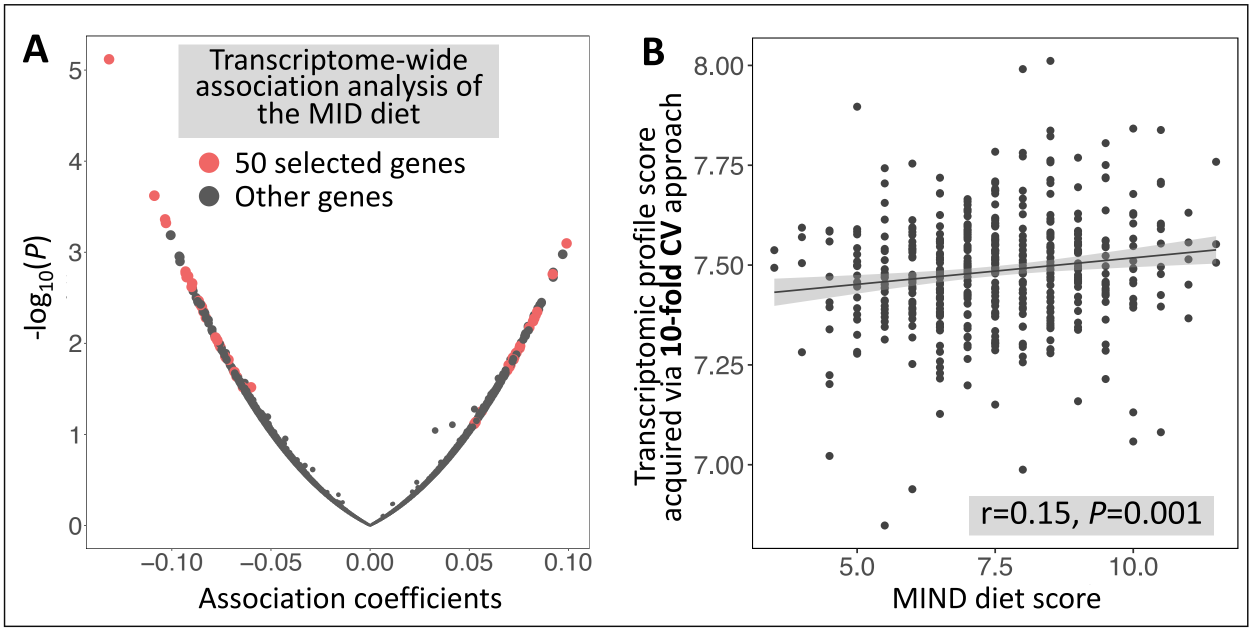

**Supplementary Figure S2. Sensitivity analysis for associations between the DLPFC expressions of the 50 selected genes with the MIND diet score.** Linear regressions were used to regress inverse normal transformed expression levels of each gene on the MIND diet score; a multivariable (MV) model was adjusted for age at death, sex, education years, total energy intake, and years from the time of FFQ to time of death. Association coefficients for the 50 genes from MV model were compared with those from models **A)** further adjusting for body mass index (BMI), physical activity, and smoking status at the time of the FFQ; **B)** further excluding individuals who were diagnosed with dementia at the time of FFQ; and further adjusting for proportions of cell types (neurons, astrocytes, microglia, oligodendrocytes) estimated based on cell-type specific gene markers derived from **C)** human brain cell-sorted RNA-seq data ("Darmanis") and from **D)** human brain single-cell RNA-seq data ("Zhang").

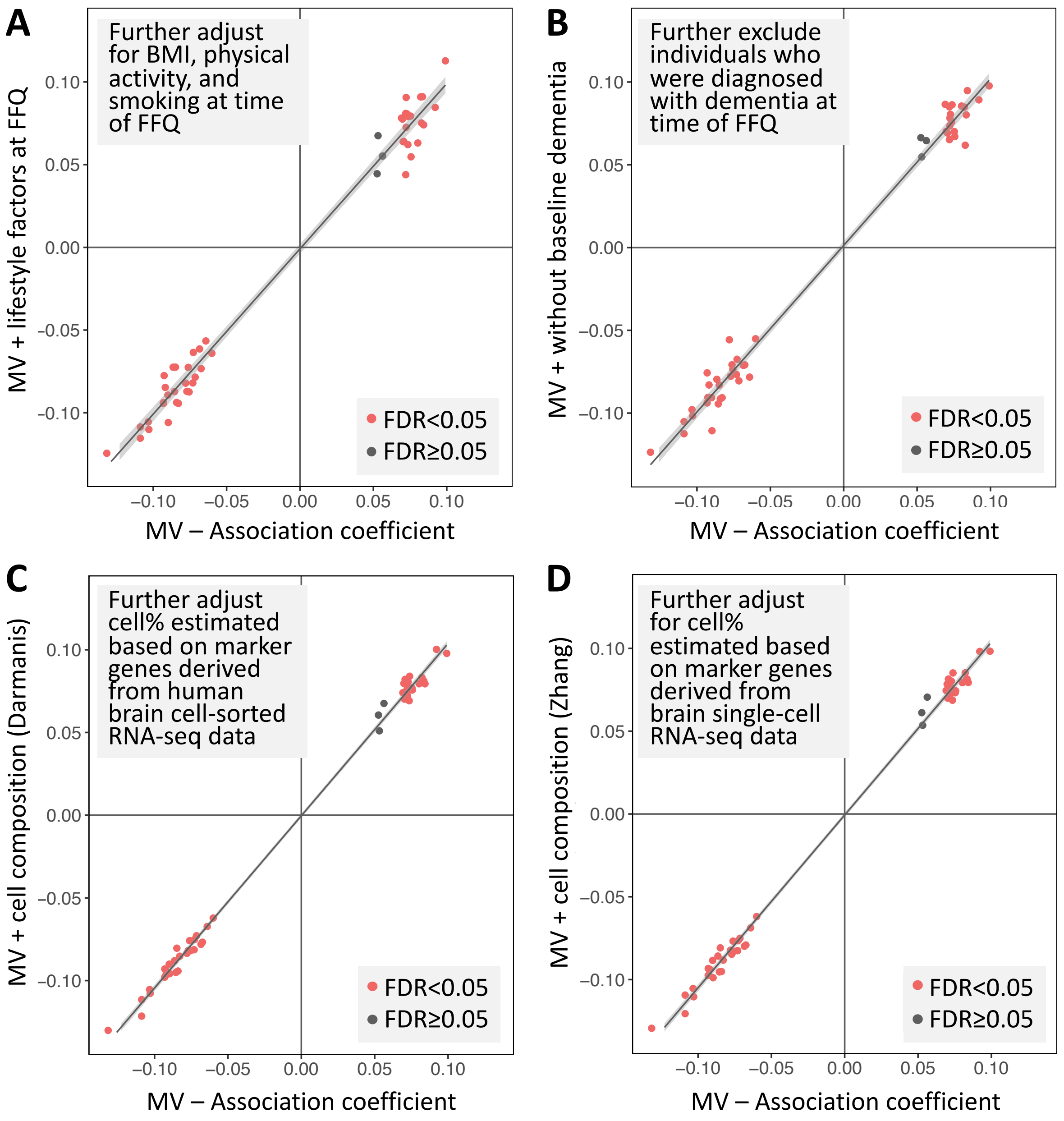

**Supplementary Figure S3. Associations between the DLPFC expression levels of the 50 selected genes with cognitive trajectory (A) and dementia risk (B), comparing results from the independent set (y-axis) with those from the combined analysis of all participants (x-axis).** Associations were adjusted for age at death, sex, education, and cohort.

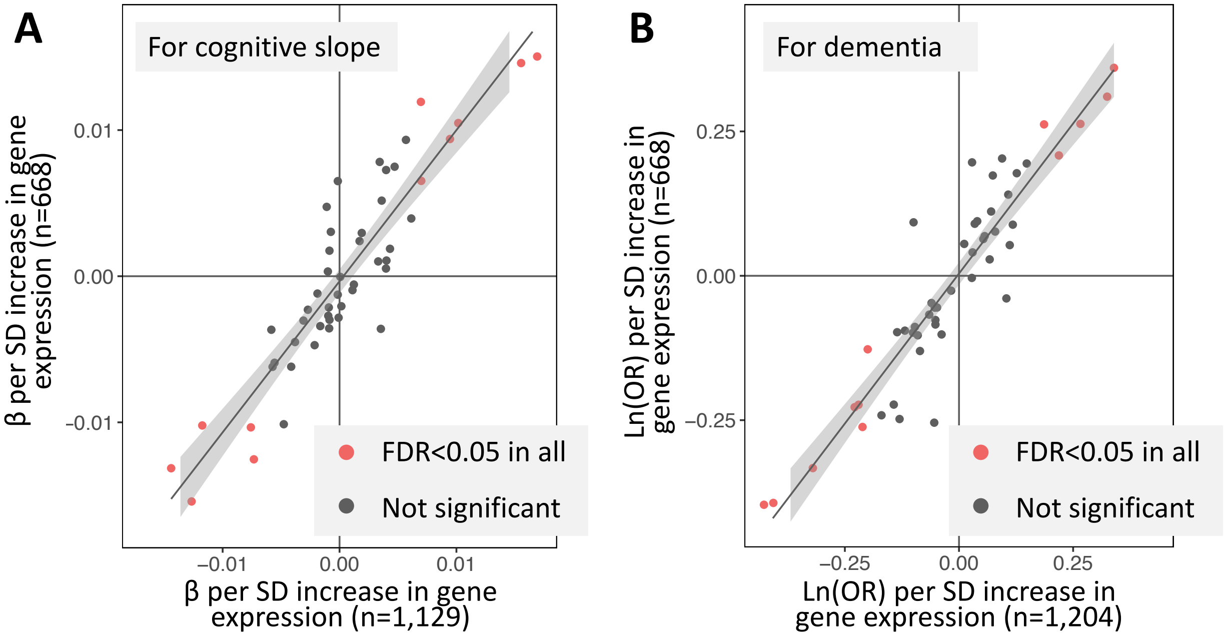

**Supplementary Figure S4. Mediation effects of selected genes in the associations between the MIND diet score and cognitive outcomes. A.** for the association between MIND diet and cognitive slope, we examined the mediation effect of 6 genes showing significant association with both the MIND diet and cognitive slope in a direction consistent with the epidemiological association between the MIND diet and cognitive slope. The multi-biomarker mediation analysis identified 3 genes showing the independent mediating effect that correspond to 30% of the total association effect. **B.** we conducted a similar analysis to examine the mediation effect of 8 genes which were significantly associated with both MIND diet and dementia. The multi-biomarker mediation analysis identified 5 genes showing the independent mediating effect that correspond to 41% of the total association effect. Analyses were conducted among participants with dietary and RNA-Seq data, and were adjusted for age at death, sex, and education years.

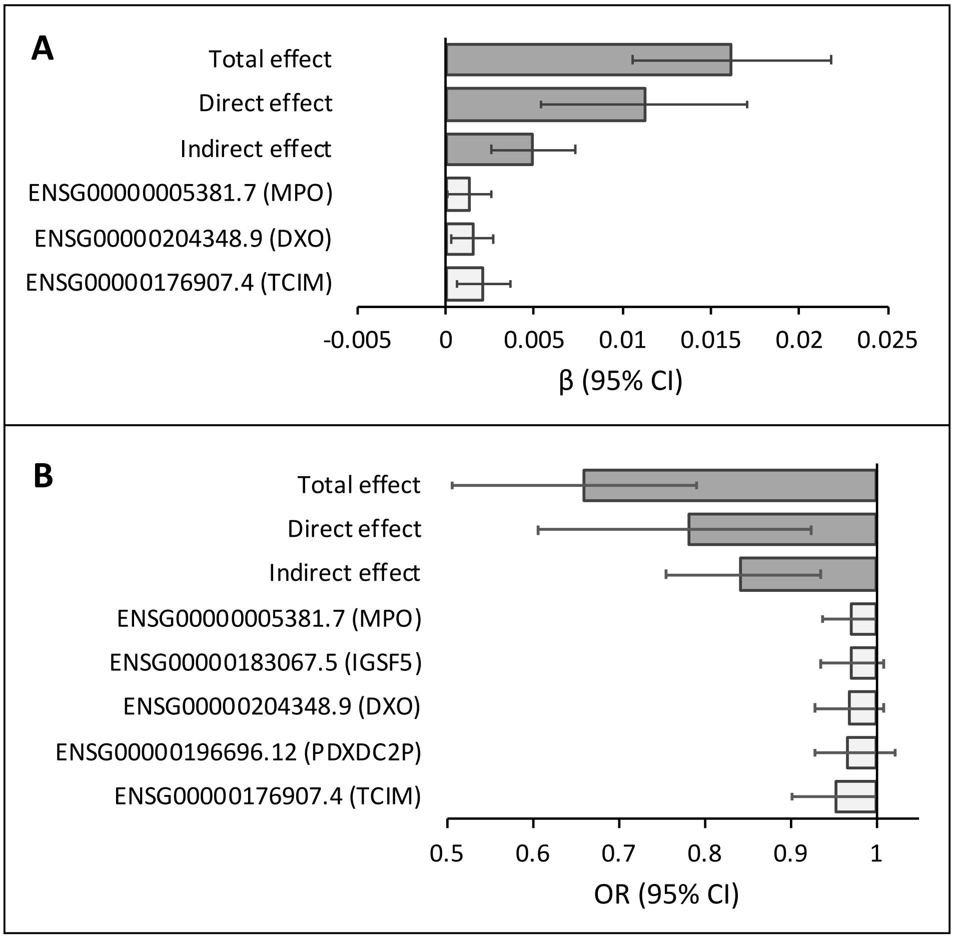

**Supplementary Figure S5. Expression levels of genes in the transcriptomic profile of MIND diet by cell types, based on DLPFC single-nuclei RNA-Seq data in 424 ROSMAP participants.** Of the 50 genes in the transcriptomic profile, we showed the cell type-specific expression levels of 32 genes that reached minimal expression threshold in at least one cell type after quality control.

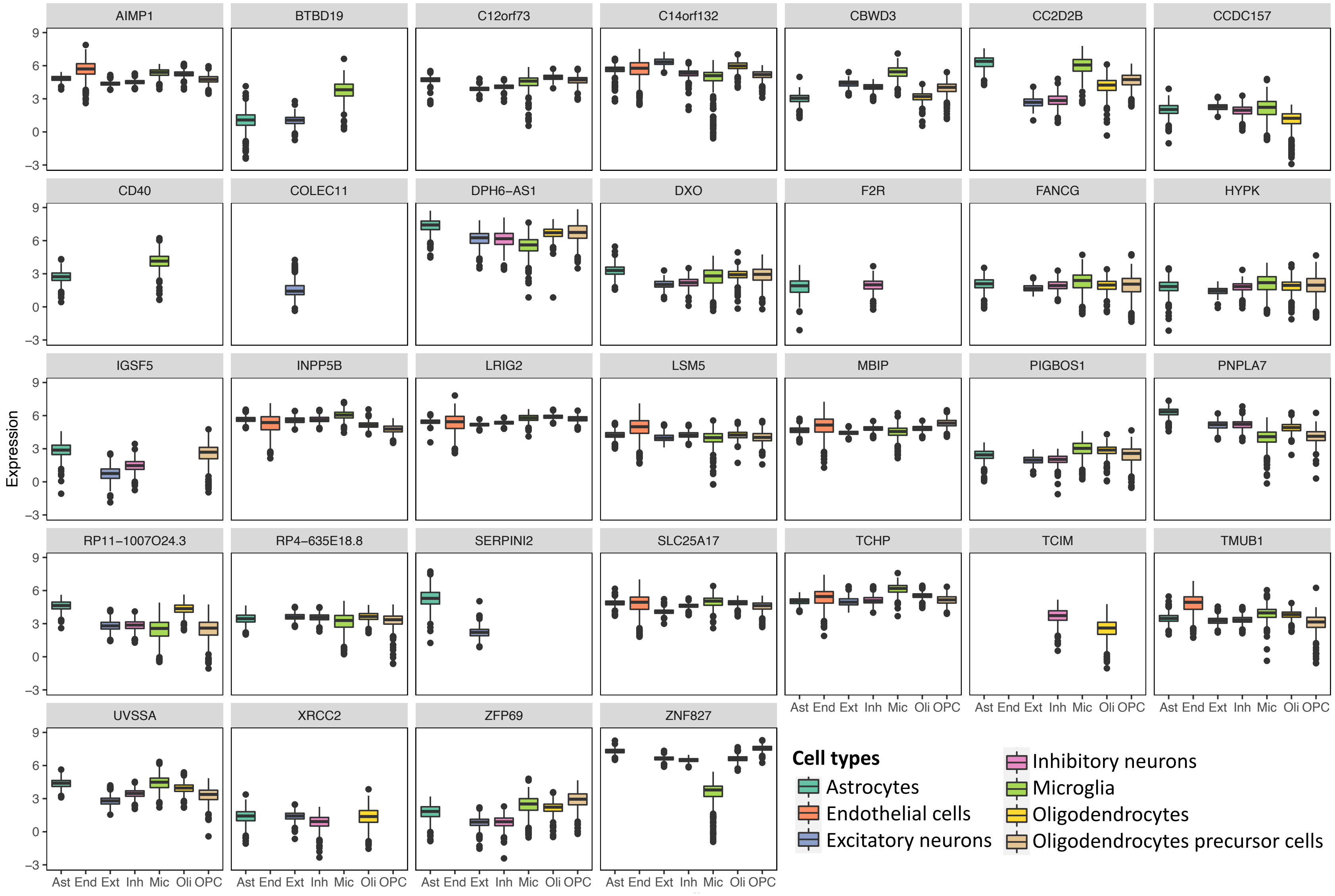

**Supplementary Figure S6. Associations between cell type-specific expression levels of selected genes with cognitive trajectory (A) and dementia (B), in 424 ROSMAP participants with DLPFC single-nuclei RNA-Seq and clinical data.** For genes significantly associated with cognitive slope and dementia in Figure 2, we examined their cell type-specific expressions in association with cognitive outcomes, adjusting for age at death, sex, years of education, and cohort. Colors of the cells represent association coefficients, with grey color (and ×) indicating that the genes did not reach minimal expression threshold in the corresponding cell types. * *P*<0.05; ** FDR<0.05 (multiple testing correction for 11 genes selected for cognitive slope analysis and 12 genes for dementia analysis).

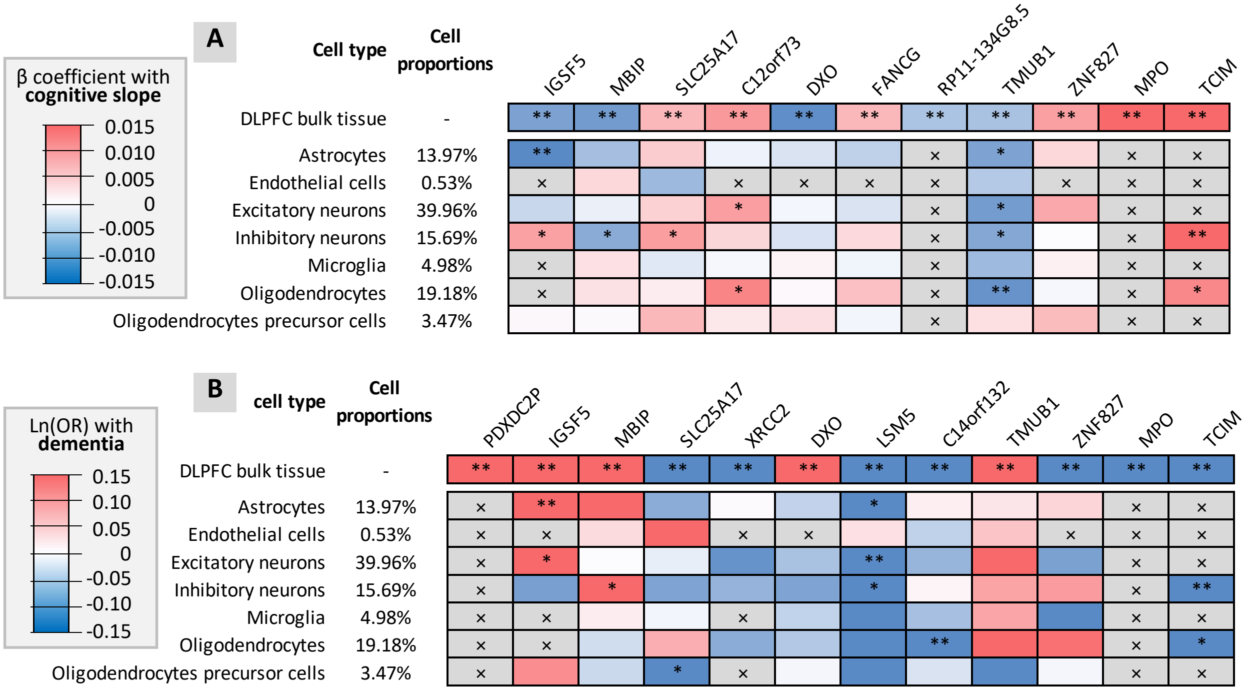
